# Ketamine’s pharmacokinetic and pharmacodynamic interactions with anticonvulsants, and their implications for psychiatry: A scoping review protocol

**DOI:** 10.1101/2023.02.09.23285702

**Authors:** Hans-Christian Stein, Rebecca Strawbridge, Daniel Silman, Pietro Carmellini, Allan H. Young, Mario F. Juruena

**Affiliations:** Department of Health Sciences, Università degli Studi di Milano, Milan, Italy; Department of Psychological Medicine, Institute of Psychiatry, Psychology and Neuroscience, King’s College London, London, UK; Department of Mental Health and Sensory Organs, University of Siena Medical Center, Italy

## Abstract

Considering the ever-increasing use of ketamine as a rapid-acting and efficacious agent for difficult-to-treat affective disorders, it is paramount that clinicians understand pharmacological interactions between ketamine and other drugs commonly used in unipolar and bipolar depression. Anticonvulsants appear of particular interest in this context, as some (valproate, lamotrigine, carbamazepine, pregabalin, gabapentin) are widely employed as mood-stabilisers and/or anxiolytics, while others are still commonly encountered due to the significant comorbidity between affective disorders and epilepsy.

This is the protocol for a scoping review aiming to comprehensively collect evidence regarding pharmacodynamic and pharmacokinetic interactions between ketamine (and its enantiomers) and anticonvulsants. Our review was designed in accordance with the PRISMA Extension for Scoping Reviews (PRISMA-ScR). PubMed, Embase, PsycINFO, ProQuest and ClinicalTrials.gov will be searched for relevant papers. Primary and non-primary research of any design will be included to ensure comprehensiveness. All data will be synthetised and presented in narrative form. We anticipate that our results will provide useful insight for clinicians and highlight gaps in knowledge that may be explored in future research.

## BACKGROUND

Over the past few decades, ketamine has established itself as an effective and rapid-acting agent for difficult-to-treat unipolar and bipolar depression^[1-4]^. An ever-growing body of literature demonstrates robust antidepressant properties of both (R-S)-ketamine (oral/parenteral) and its enantiomer S-ketamine, approved in 2019 as a nasal spray by the FDA and EMA for treatment-resistant depression^[2-6]^. Although its mechanism of action is multifaceted and involves several neurochemical pathways (i.e. GSK3, mTOR, BDNF), ketamine exerts its antidepressant effects primarily as a NMDA receptor antagonist, by blocking the tonic glutamatergic stimulation of GABAergic interneurons and thus eventually enhancing AMPAR-related glutamate signalling^[7]^.

As the use of ketamine for severe mood disorders is growing increasingly popular, especially for patients with long courses of illness and complex pharmacological treatments, it is imperative that psychiatrists are aware of pharmacological interactions and combined effects of ketamine and other drugs commonly encountered in clinical practice. Antiepileptics appear to be of particular interest in this regard, both for the involvement of GABAergic and glutamatergic circuits in their mechanism of action^[8,9]^ and for their wide scope of use. Indeed, molecules like valproate, lamotrigine, carbamazepine, pregabalin and gabapentin are commonly prescribed in affective disorders as mood stabilisers or/and anxiolytics. Other antiepileptic medication, though not having an indication in psychiatric conditions, are still commonly encountered in clinical psychiatry since affective disorders are a most common comorbidity in epilepsy^[10]^.

However, solid evidence regarding interactions between anticonvulsants and ketamine appears exiguous and dispersed. We designed this scoping review in order to comprehensively synthetise all available evidence in this field, identify relevant gaps in knowledge, and provide insight useful for clinical practice.

## OBJECTIVE

To review and summarise available evidence of pharmacokinetic and pharmacodynamic interactions between ketamine (R,S-ketamine, S-ketamine or R-ketamine) and antiepileptic drugs.

## SEARCH STRATEGY AND INFORMATION SOURCES

A scoping approach was chosen in lieu of a systematic review design due to the breadth of our reseach question and our aim to paint a complete picture of available literature.

We will search several sources for published and unpublished records using relevant keywords and thesauri: PubMed, Embase, PsycINFO, ProQuest and ClinicalTrials.gov. No time or language restriction will be applied. Reference lists of resulting records (included studies and related reviews) will be inspected, to retrieve any additional relevant studies.

### Search strategy for PubMed

(“ketamine”[MeSH Terms] OR “ketamine”[Title/Abstract] OR “esketamine”[Title/Abstract] OR “arketamine”[Title/Abstract] OR “S-ketamine”[Title/Abstract] OR “r-ketamine”[Title/Abstract]) AND (“valproate”[Title/Abstract] OR “valproic”[Title/Abstract] OR “divalproex”[Title/Abstract] OR “lamotrigine”[Title/Abstract] OR “carbamazepine”[Title/Abstract] OR “oxcarbazepine”[Title/Abstract] OR “pregabalin”[Title/Abstract] OR “gabapentin”[Title/Abstract] OR “topiramate”[Title/Abstract] OR “clonazepam”[Title/Abstract] OR “clobazam”[Title/Abstract] OR “clorazepate”[Title/Abstract] OR “phenytoin”[Title/Abstract] OR “mephenytoin”[Title/Abstract] OR “phenobarbital”[Title/Abstract] OR “felbamate”[Title/Abstract] OR “lacosamide”[Title/Abstract] OR “zonisamide”[Title/Abstract] OR “rufinamide”[Title/Abstract]OR “vigabatrin”[Title/Abstract] OR “tiagabine”[Title/Abstract] OR “primidone”[Title/Abstract] OR “levetiracetam”[Title/Abstract] OR “anticonvuls*”[Title/Abstract] OR “anti-convuls*”[Title/Abstract] OR “antiepilep*”[Title/Abstract] OR “anti-epilep*”[Title/Abstract] OR “antiseizure*”[Title/Abstract] OR “anti-seizure*”[Title/Abstract])

The same search strategy will be used for the other databases after syntax adaptation.

## STUDY SELECTION

All qualitative, quantitative or mixed-methods studies presenting findings relevant to any possible interaction between ketamine (R,S-ketamine, S-ketamine or R-ketamine) and maintenance anticonvulsants, both administered at any dosage and duration, via any route and for any indication or participant cohort will be included. Rescue anti-seizure drugs i.e. benzodiazepines other than clonazepam, clobazam and clorazepate will not be considered. RCTs, observational studies, case reports and case series involving human subjects of all ages will be included, as well as any eligible pre-clinical studies or animal studies. Grey literature will be included. Systematic reviews and literature reviews will be excluded.

The following criteria will be applied in regard to concomitant medications:

### Interventional studies:□

- In single-arm trials,□standardised drug interventions concurrent with ketamine/anticonvulsants are NOT allowed.
- In two+ arm trials, standardised drug interventions concurrent with ketamine/anticonvulsants are allowed only if administered to all arms OR if at least one ketamine/anticonvulsant arm is unaffected.
- In both cases, the presence/absence of non-standardised concomitant drug treatments must be stated and, if present, they must be described.

### Case reports

- Presence/absence of concomitant drug treatments must be stated and, if present, they must be described.
- Studies are excluded if concomitant drug treatments have been initiated or titrated up within 24 hours of outcome measurement.□

### Naturalistic studies

- For descriptive, single-group studies, exposure to specific drug treatments concurrent with ketamine/ anticonvulsants is NOT allowed, except unless the manuscript states a rate of□<10% participants.
- In comparative multiple-group studies, exposure to specific drug treatments□concurrent with ketamine/ anticonvulsants□is allowed unless the manuscript explicitly states any between-group difference in concomitant interventions, although if a rate of□<10% of participants is stated, this is allowed.

Two authors will independently perform the abstract screening of the titles retrieved by the search. Articles included by at least one author will be eligible for full-text screening. Once the full articles are obtained, the full-text screening will be carried out independently by two investigators. Disagreement will be resolved by consensus or by consultation with a senior member of the review team.

## DATA SELECTION AND CHARTING

Two investigators will systematically extract relevant data from each included study (including study characteristics, methodology and results) by using a data extraction tool designed for this study. Differences will be discussed and resolved by consensus or by consultation with a senior member of the review team. Should further information or data be deemed necessary, an attempt to contact the original authors will be made.

## DATA SUMMARY AND SYNTHESIS OF RESULTS

Results of the study selection and data extraction will be presented in both narrative format and visual representation. The screening process will be documented using a PRISMA flow diagram. Reasons for exclusion will be provided. Findings will be summarised for each ketamine-anticonvulsant pairing and clinical implications will be discussed.

## PROTOCOL AND REGISTRATION

This protocol was drafted in accordance to the PRISMA Extension for Scoping Reviews (PRISMA-ScR)^[11]^ and the JBI Manual for Evidence Synthesis^[12]^. The protocol will be registered and made available online in the medRxiv.org database.

## Data Availability

All data produced in the present work are contained in the manuscript.

